# Evaluating Clinical Outcomes in Patients Being Treated Exclusively via Telepsychiatry

**DOI:** 10.1101/2023.09.27.23296225

**Authors:** Cheryl Person, Nicky O’Connor, Lucy Koehler, Kartik Venkatachalam, Georgia Gaveras

## Abstract

**Background:** Depression and anxiety are highly prevalent conditions in the US. Despite the availability of suitable therapeutic options, limited access to high quality psychiatrists represents a major barrier to treatment. Although telepsychiatry has the potential to improve access to psychiatrists, treatment efficacy in the telepsychiatry model remains unclear.

**Objectives:** Our primary objective was to determine whether there was a clinically meaningful change in one of two validated outcome measures of depression and anxiety —the Patient Health Questionnaire-8 (PHQ8) or the Generalized Anxiety Disorder Questionnaire-7 (GAD7) — after receiving at least 8-weeks of treatment in an outpatient telepsychiatry setting.

**Methods:** Treatment-seeking patients enrolled in a large outpatient telepsychiatry service that accepts commercial insurance. All analyzed patients completed GAD7 and PHQ8 prior to their first appointment, and at least once after 8 weeks of treatment. Treatments included comprehensive diagnostic evaluation, supportive psychotherapy, and medication management.

**Results:** 1826 treatment-seeking patients were evaluated for clinically meaningful changes in GAD7 and PHQ8 scores during treatment. Mean treatment duration was 103 days (SD =34). At baseline, 58.8% and 60% of patients exhibited at least moderate anxiety and depression, respectively. In response to treatment, mean change for GAD7 was -6.71 (95% CI -7.03, -6.40) and for PHQ8 was -6.85 (95% CI -7.18, -6.52). Patients with at least moderate symptoms at baseline, showed 45.7% reductions in GAD7 scores and 43.1% reductions in PHQ8. Effect sizes for GAD7 and PHQ8, as measured by Cohen’s *d* for paired samples, were *d* = 1.30 (*P*<.001) and 1.23 (*P*<.001) respectively.

Changes in GAD7 and PHQ8 scores correlated with the type of insurance held by the patients. Greatest reductions in scores were observed among patients with commercial insurance (45.0% and 43.9% reductions in GAD7 and PHQ8, respectively). Although Medicare patients did exhibit statistically significant reductions in GAD7 and PHQ8 scores from baseline, these improvements were attenuated compared to those in patients with commercial insurance (29.2% and 27.6% reduction in GAD7 and PHQ8, respectively). Pairwise comparison tests revealed significant differences in treatment responses in patients with Medicare vs. commercial insurance (*P* < .001). Responses were independent of patient geographic classification (urban vs rural; *P* = .475 for GAD7 and *P* = .065 for PHQ8). The findings that treatment efficacy was comparable among rural and urban patients indicated that telepsychiatry is a promising approach to overcome treatment disparities that stem from geographical constraints.

**Conclusion:** In this large retrospective data analysis of treatment seeking patients utilizing a telepsychiatry platform, we found robust and clinically significant improvement in depression and anxiety symptoms during treatment. The results provide further evidence that telepsychiatry is highly effective and has the potential to improve access to psychiatric care.

## Introduction

Depression and anxiety are the two most common mental health conditions in the US and worldwide, ranking 2^nd^ and 8^th^, respectively, in the list of causes for disability [1]. The chronicity and prevalence of these illnesses have an enormous economic impact and have worsened the quality of life in countries across the globe. A recent health economics review found that depression alone accounts for roughly 16% of the loss of work-related productivity [2]. Those who suffer from depression and anxiety have worse health outcomes if left untreated [3–5].

Although effective treatment is available for both these chronic conditions, access to these treatments can be a challenge, which has led to significant gaps in treatment [6–8]. Difficulties in access to treatment can stem from physical distances from a treatment site, difficulties with arranging for time away from home or work to attend clinical appointments, prohibitive treatment costs, and the paucity of specialist care [9,10]. Remarkably, 27% of the counties in the US are characterized by the complete absence of psychiatrists [11]. Rural residents are noteworthy for having limited access to in person psychiatric care [9,11]. Even more troubling, it appears that issues of access occur independently of the patients’ health insurance status. A recent survey found that 31% of US adults reported the inability to access mental health services despite being insured [12]. Further, the medical specialty least likely to participate in health insurance plans is psychiatry [13]. Consequently, the total cost for outpatient psychiatric care is often not mitigated by health insurance. Not only is outpatient psychiatric care without insurance often cost prohibitive, potential paucity of in-network psychiatrists may further delay access to treatment. These delays in access to mental health treatment are burdensome to patients, and frequently worsen other associated medical conditions [14,15].

The aforementioned obstacles are especially disconcerting given the efficacy of treatments for both depression and anxiety. Antidepressants and psychotherapy have long been recognized as being beneficial for treating these conditions [16,17]. Given that modern technology permits the delivery of evidence-based treatments remotely, and that psychiatry as a practice, does not require the presence of patients and psychiatrists in the same physical space, telepsychiatry is a valuable tool for overcoming these persistent barriers. Indeed, the recent easing of regulatory obstacles to the implementation of telepsychiatry platforms has led to a vast expansion in the ease of access to mental health services. Encouragingly, a growing body of literature indicates that patients receiving telehealth treatments exhibit improvements that are comparable to those seen in patients receiving in-person care [18–20]. Others have demonstrated that a combination of flexibly implemented psychotherapy and medication management is effective for management of anxiety disorders [21]. Presently, the potential efficacy of telepsychiatry services for patients who elect a fully virtual telepsychiatry service remains unknown.

In this study, we examined the outcomes of a large private outpatient telepsychiatry practice. Talkiatry is a fully virtual, nationwide outpatient private psychiatry practice that accepts commercial health insurance. This practice employs over 300 psychiatrists and more than 50 therapists across the US. Although virtual, Talkiatry utilizes the traditional psychiatric model of care — an initial comprehensive evaluation, followed by the development of treatment plan that revolves around patient preferences. This type of patient-centric approach comprised of partnerships between physicians and patients that are built on the foundation of effective communication and health promotion has been found to improve medical outcomes and forms the basis of effective outpatient psychiatric care [22].

The concept of therapeutic alliance— the collaborative relationship between the patient and physician — is another core element of psychiatric care that has been described as one of the most valuable aspects of treatment effectiveness with strong positive impact on outcomes in psychotherapy [23–25]. In fact, a patient’s adherence to treatment is often motivated by a strong therapeutic alliance [26]. Further, recent studies have suggested that therapeutic alliance is the mediator of change in psychotherapy studies across a broad range of modalities and conditions [27]. The efficacy of therapeutic alliance depends on the ability of the physician to convey empathy and friendliness, to understand a patient’s unique set of motivations and interests, and to implement an individualized treatment that is personalized to that patient’s preferences [28] Not surprisingly, it takes time to build a therapeutic alliance. Therefore, the establishment of therapeutic alliance is difficult in the common and increasingly dominant 15-minute medication checks that characterize current psychiatric practice [29,30]. Here, we examined whether the Talkiatry telepsychiatry model, which has time set aside specifically for the establishment of a strong therapeutic alliance, led to improved clinical outcomes for patients with anxiety or depression.

## Objectives

The primary objective of this study was to determine whether there was a clinically-meaningful change in one of two validated outcome measures — the Patient Health Questionnaire-8 (PHQ8) or the Generalized Anxiety Disorder Questionnaire (GAD7) — after receiving treatment in the outpatient telepsychiatry practice for a minimum of 8-weeks. This retrospective data analysis reflects real-world treatment conditions where the combination of therapy and medication management remains the primary modality of treatment. Our secondary objectives were to determine if patient demographic characteristics (sex, urban/rural, initial GAD7/PHQ8 severity score, age, and insurance status) correlated with statistically significant changes in treatment outcomes.

## Methods

### Study cohort and treatment strategy

The study population was comprised of treatment-seeking individuals who had enrolled in an outpatient telepsychiatry practice. Patients sought enrollment in services through several commercially available booking platforms or the company website and required the completion of a brief online questionnaire. Enrollment was restricted to individuals who did not present with primary psychotic disorder, primary substance use disorder, recent psychiatric hospitalization, current suicidal ideation, and insurance types not accepted by the practice. Once screening was completed, patients were provided several psychiatrists’ profiles to help them choose the provider that met their unique preferences (gender, condition specialty, availability outside of business hours, etc.). From that curated group, each patient selected a psychiatrist and scheduled a new patient evaluation. All patients were requested to complete GAD7 and PHQ8 test up to seven days prior to their first appointment, and approximately every month thereafter. Enrolled patients were 18 years of age or older and had at least one baseline measure and one follow-up measure that were completed during the specified timeframe. The follow-up measure was included if it occurred at least eight weeks after the initial measure. If a patient completed multiple measures, the last completed measure was selected for analysis. This study was granted an exemption by an external institutional review board.

Therapy and medication management were provided in an audiovisual format that utilized a commercially available telehealth platform. Psychiatrists conducted comprehensive psychiatric evaluations, and in collaboration with the patient, developed appropriate treatment plans. Ongoing treatments, which included supportive psychotherapy, developing therapeutic alliance, and medication management, were embedded within the appointments.

### Measures

The PHQ8 is an 8-item self-reporting questionnaire that quantifies the symptoms of depression, and measures progress in patient outcomes over time [31]. The scale performs similarly to the PHQ9 and is routinely used in telehealth research and clinical practice settings [32]. For PHQ8 the standard cutoff scores were utilized to categorize patients into minimal (0-4), mild (5-9), moderate (10-14), moderately severe (15-19), and severe (20-24). The GAD7 is a 7-item self-reporting questionnaire that quantifies symptoms of generalized anxiety disorder, and track progress of patient outcomes over time [33]. For GAD7 the standard cutoff scores were utilized to categorize patients into minimal (0-4), mild (5-9), moderate (10-14), and severe (15-21). Initial PHQ8 and GAD7 scores were collected within 7-days of the first appointment, and at approximately one-month intervals during treatment. Demographic variables that were analyzed included age, sex, type of health insurance (commercial insurance, Medicare, or self-pay), and urban/rural classification. Self-pay was assigned to patients who were initially insured but changed or lost insurance during the course of treatment.

### Statistical Analysis

Primary outcomes of interest in this study were anxiety and depressive symptoms measured by GAD7 and PHQ8, respectively, assessed at the last visit. Changes in GAD7 and PHQ8 scores were calculated by subtracting the last scores from baseline scores for each patient. In order to examine clinically-relevant samples, only patients with at least mild symptoms at baseline (i.e., GAD7≥ 5, PHQ8 ≥ 5) were included in the analyses. We use paired sample Wilcoxon tests to examine whether statistically significant changes in GAD7 and PHQ8 were observed over time (from baseline to last visit) among all patients. Effect sizes for paired sample t-tests were also calculated using the standard deviation of the differences. These analyses were repeated with subsamples including patients with at least moderate baseline symptoms (i.e., baseline GAD7 score ≥ 10 and baseline PHQ8 score ≥ 10).

We examined whether changes in anxiety and depressive symptom scores varied by sex, age group (18-24 years, 25-64 years, and 65+ years), baseline symptoms-severity category, insurance type (i.e., commercial insurance, Medicare, self-pay), and geographical classification (i.e., urban vs. rural) using Wilcoxon, Kruskal-Wallis or Chi-square tests. Binary outcomes indicating patients that showed clinically-significant symptom improvement were, 1 for patients with 50% or greater symptoms improvement, and 0 for patients not at goal. For these outcomes, we chose to include only those patients with at least moderate baseline symptoms (i.e., baseline GAD7≥ 10, PHQ8 score ≥ 10).

We determined the fraction of patients with > 50% symptom improvement, including those that exhibited remission (final score <5). Using the Chi-square test, we asked whether the fraction of patients with symptom improvement varied as a function of the category of baseline symptom severity. In these analyses, we included patients with at least moderate baseline symptoms because GAD7 and PHQ8 scores ≥10 represent a high probability of clinically significant symptoms [31]. Statistical analyses were performed using R (R core team 2021 https://www.R-project.org/) and Prism (Version 10.0.02; GraphPad).

## Results

The demographic and baseline characteristics of the patients are presented in Table 1. During the study timeframe, a total of 6465 patients enrolled in outpatient care, and had a baseline measure available. Of those, 1826 participants, representing 39% of the initial sample completed a second measure and were included in the analysis. A total of 1826 participants had data available for analyses and had a mean age of 39.4 years (SD=13.5, range = 18-89). Of those who reported sex (n=1479, 19% did not report sex), 1084 (73.3%) were female. The mean number of days between baseline and last visit was 103 days (SD = 34). Of those 1826, 1602 (87.7%) patients reported at least mild anxiety symptoms (GAD7 ≥ 5) while 1572 (86.1%) patients reported at least mild depressive symptoms (PHQ8 score ≥ 5) at baseline. In addition, 58.8% of sample reported at least moderate anxiety symptoms and 60% of sample reported at least moderate depressive symptoms. Of the original sample, a total of 4639 patients who did not complete the second measure. Compared to patients who were non-completers, those who completed repeated measures were older, more likely to be insured with Medicare, and more likely to be classified as rural. Importantly, they did not differ on baseline severity for either depressive or anxiety symptoms (Table 1).

**Table 1.** Demographic and Baseline Characteristics.

| Baseline Variables |  | Patients with<br>repeat measures<br>Mean (SD)/n<br>(%) | Patients with<br>one measure<br>Mean (SD)/n<br>(%) | *P*-value* |
| --- | --- | --- | --- | --- |
| Sex |  |  |  | .054 |
|  | Female | 1084 (59.36%) | 2620 (56.48%) |  |
|  | Male | 395 (21.63%) | 1086 (23.41%) |  |
|  | Undisclosed | 347 (19.00%) | 933 (20.11%) |  |
| Age |  | 39.38 (13.53) | 33.32 (10.05) | <.0001 |
| Insurance Type |  |  |  | <.0001 |
|  | Commercial Insurance | 1618 (88.61%) | 4409 (95.04%) |  |
|  | Medicare | 138 (7.56%) | 84 (1.81%) |  |
|  | Self-Pay | 70 (3.83%) | 146 (3.15%) |  |
| Baseline GAD7 |  |  |  | .90 |
|  | Minimal anxiety | 224 (12.27%) | 544 (11.73%) |  |
|  | Mild anxiety | 528 (28.92%) | 1332 (28.71%) |  |
|  | Moderate anxiety | 541 (29.63%) | 1376 (29.66%) |  |
|  | Severe anxiety | 533 (29.19%) | 1387 (29.9%) |  |
| Baseline PHQ8 |  |  |  | .49 |
|  | Minimal depression | 254 (13.91%) | 567 (12.22%) |  |
|  | Mild depression | 475 (26.01%) | 1260 (27.16%) |  |
|  | Moderate depression | 530 (29.03%) | 1383 (29.81%) |  |
|  | Moderately severe depression | 394 (31.58%) | 1032 (22.25%) |  |
|  | Severe depression | 173 (9.47%) | 397 (8.56%) |  |
| Geographic Classification |  |  |  | .01 |
|  | Urban | 1710 (93.65%) | 4420 (95.28%) |  |
|  | Rural | 116 (6.35%) | 218 (4.7%) |  |
| Number of days between visits |  | 102.63 (34.15) | n/a |  |

Abbreviations: PHQ8 Patient Health Questionnaire-8, GAD7 Generalized Anxiety Disorder Questionnaire-7
\**P*-values obtained from chi-square tests comparing group that completed repeated measurements and group that completed one measurement. n/a= not available as second measurement not completed

### Changes in Anxiety and Depressive Symptoms

Results from paired sample Wilcoxon tests showed that among patients with at least mild baseline symptoms (GAD7/PHQ8 ≥ 5), both anxiety (GAD7 scores) and depressive symptoms (PHQ8 scores) decreased significantly from the first to last visit. The effect sizes for GAD7 and PHQ8, as measured by Cohen’s *d* for paired samples, were *d* = 1.05 (*p*<.001) and 0.98 (*p*<.001), respectively. These findings indicatied large effects (43.7% reductions in GAD7, 42.7% reductions in PHQ8). Furthermore, these findings were replicated when the analyses were conducted with the subsample of patients that exhibited at least moderate baseline symptoms (GAD7/PHQ8 ≥ 10). We observed larger effect sizes of *d* = 1.30 (p<.001) for GAD7 (45.7% reductions) and *d* = 1.23 (p<.001) for PHQ8 (45.1% reductions) in the subsample. These data indicated that greater reductions in anxiety and depressive symptoms occurred among patients with more severe symptoms at baseline (Table 2).

**Table 2.**

| Measure | Sample | Responses,<br>n | Baseline<br>Score,<br>Mean (SD) | Last<br>Score,<br>Mean (SD) | Changes<br>from<br>Baseline, % | Mean Difference<br>(95% CI) | Effect Size<br>(Cohen's d) | Baseline<br>Score,<br>Median | Last<br>Score,<br>Median | Paired<br>Sample<br>Wilcoxon<br>Test | P-value |
| --- | --- | --- | --- | --- | --- | --- | --- | --- | --- | --- | --- |
| GAD7 | Baseline GAD7 $\geq 5$ | 1602 | 12.20 (4.55) | 6.87 (4.82) | 43.69 | -5.34 (-5.59, -5.09) | 1.05 | 12 | 6 | V=1099198 | < .001 |
| | Baseline GAD7 $\geq 10$ | 1074 | 14.72 (3.27) | 8.00 (5.00) | 45.65 | -6.71 (-7.03, -6.40) | 1.30 | 14 | 7 | V=1039814 | < .001 |
| PHQ8 | Baseline PHQ8 $\geq 5$ | 1572 | 12.78 (4.90) | 7.33 (5.47) | 42.64 | -5.45 (-5.73, -5.18) | 0.98 | 12 | 6 | V=519504 | < .001 |
| | Baseline PHQ8 $\geq 10$ | 1097 | 15.22 (3.74) | 8.37 (5.69) | 45.01 | -6.85 (-7.18, -6.52) | 1.23 | 15 | 8 | V=482402 | < .001 |

Note: Patients with at least mild baseline symptoms (baseline GAD7/PHQ8 $\geq 5$ ) are included in the analyses.

#### Sex

Kruskal-Wallis tests showed that the overall fraction of patients with 50% or greater reductions in either GAD7 or PHQ8 did not differ by sex (GAD7, *P* = .33; PHQ8, *P* = .10).

#### Baseline Symptom Severity

Using Kruskal-Wallis tests to examine whether change scores in GAD7 and PHQ8 differed across baseline symptom severity category, we found greater reductions among those with more severe baseline symptoms. The greatest reductions were among patients with severe baseline symptoms (GAD7 ≥ 15, PHQ8 ≥ 20). The smallest reductions were among patients with mild baseline symptoms (GAD7/PHQ8 = 5-9) (Table 3). Pairwise comparisons using Wilcoxon test with continuity correction revealed that significant differences in change scores in GAD7 and PHQ8 were observed between all comparison pairs (all *P* <.01). Therefore, groups with more severe baseline symptoms showed significantly greater reductions in anxiety and depressive symptoms compared to any of the groups with lower baseline severity.

**Table 3.**

| Measure | Baseline Severity | Responses,<br>n (%) | Baseline<br>Score,<br>Mean (SD) | Last Score,<br>Mean (SD) | Changes<br>from<br>Baseline, % | Mean Difference<br>(95% CI) | Baseline<br>Score,<br>Median | Last<br>Score<br>Median | Kruskal -<br>Wallis | P-value |
| --- | --- | --- | --- | --- | --- | --- | --- | --- | --- | --- |
| GAD7 | Mild anxiety (5-9) | 528 (32.96%) | 7.09 (1.39) | 4.55 (3.42) | 35.83 | -2.54 (-2.84, -0.24) | 7 | 4 | 336.85 (df=2) | < .001 |
|  | Moderate anxiety (10-14) | 541 (33.77%) | 11.91 (1.37) | 6.79 (4.08) | 42.99 | -5.12 (-5.47, -4.77) | 12 | 6 |  |  |
| | Severe anxiety ($\geq 15$) | 533 (33.27%) | 17.57 (1.88) | 9.24 (5.51) | 47.41 | -8.33 (-0.81, -7.86) | 17 | 8 | | |
| PHQ8 | Mild depression (5-9) | 475 (30.22%) | 7.15 (1.34) | 4.93 (4.01) | 31.05 | -2.22 (-2.59, -1.86) | 7 | 4 | 304.6 (df = 3) | < .001 |
|  | Moderate depression (10-14) | 530 (33.72%) | 11.97 (1.40) | 6.68 (4.61) | 44.19 | -5.29 (-5.80, -4.90) | 12 | 6 |  |  |
|  | Moderately severe depression (15-19) | 394 (25.06%) | 16.91 (1.39) | 9.08 (5.94) | 46.30 | -8.83 (-8.41, -7.24) | 17 | 8 |  |  |
| | Severe depression ($\geq 20$) | 173 (11.01%) | 21.36 (1.31) | 11.95 (6.13) | 44.05 | -9.41 (-10.31, -8.51) | 21 | 12 | | |

Note: Patients with at least mild baseline symptoms (baseline GAD7/PHQ8 $\geq 5$ ) are included in the analyses.

#### Insurance Type

Insurance types (i.e., commercial insurance, Medicare, self-pay) also predicted change scores of both anxiety symptoms (GAD7) and depressive symptoms (PHQ8). Greatest reductions were observed among those with commercial insurance (45.0% reductions for GAD7, 43.9% reductions in PHQ8), followed by those with self-pay (40.8% reductions for GAD7, 40.1% reductions in PHQ8) and Medicare (29.2% reductions for GAD7, 27.6% reductions in PHQ8). Pairwise comparison tests revealed that significant differences in change scores were found between Medicare and commercial insurance (*P* < .001) and Medicare and self-pay (*P* < .01), but not commercial insurance and self-pay (*P* = .74) (Table 4).

**Table 4.**

| Measure | Insurance Type | Responses,<br>n | Baseline<br>Score,<br>Mean (SD) | Last Score,<br>Mean (SD) | Changes from<br>Baseline, % | Mean Difference<br>(95% CI) | Baseline<br>Score,<br>Median | Last<br>Score,<br>Median | Kruskal-Wallis | *P*-value |
| --- | --- | --- | --- | --- | --- | --- | --- | --- | --- | --- |
| GAD7 | Commercial Insurance | 1424 | 12.21 (4.54) | 6.72 (4.72) | 44.96 | -5.50 (-5.76, -5.23) | 12 | 6 | 14.04 (df = 2) | < .001 |
|  | Medicare | 115 | 11.61 (4.36) | 8.22 (5.41) | 29.20 | -3.39 (-4.31, -2.47) | 11 | 7 |  |  |
|  | Self-Pay | 63 | 13.11 (5.02) | 7.76 (5.44) | 40.81 | -5.35 (-6.54, -4.16) | 14 | 6 |  |  |
| PHQ8 | Commercial Insurance | 1405 | 12.76 (4.92) | 7.16 (5.40) | 43.89 | -5.60 (-5.89, -5.31) | 12 | 6 | 20.46 (df = 2) | < .001 |
|  | Medicare | 107 | 12.62 (4.930) | 9.14 (5.70) | 27.58 | -3.48 (-4.55, -2.40) | 12 | 8 |  |  |
|  | Self-Pay | 60 | 13.60 (4.43) | 8.15 (6.13) | 40.07 | -5.45 (-6.77, -4.13) | 13 | 6.5 |  |  |

Note: Patients with at least mild baseline symptoms (baseline GAD7/PHQ8 $\geq 5$ ) are included in the analyses.

#### Age Group

In the unadjusted model, age groups predicted differences in the fraction of patients with 50% or greater reductions in both GAD7 (X2 (2, N=1826) =11.44, p =.003) and PHQ8 (X2 (2, N=1826) = 9.57, p = .008).

#### Geographic Classification (Urban vs. Rural)

No differences in change scores in either GAD7 or PHQ8 were found across geographic classification (i.e., urban vs. rural) indicating that similar reductions in symptoms were observed across patients’ geographic location (Table 5).

**Table 5.**

| Measure | Geographic<br>Classification | Responses,<br>n | Baseline<br>Score,<br>Mean (SD) | Last Score,<br>Mean (SD) | Changes from<br>Baseline, % | Mean Difference<br>(95% CI) | Baseline<br>Score,<br>Median | Last<br>Score,<br>Median | Wilcoxon Rank<br>Sum Test | *P*-value |
| --- | --- | --- | --- | --- | --- | --- | --- | --- | --- | --- |
| GAD7 | Urban | 1499 | 12.19 (4.57) | 7.45 (4.88) | 38.88 | -5.36 (-5.62, -5.11) | 12 | 6 | W = 80435 (df=1) | .475 |
|  | Rural | 103 | 12.41 (4.26) | 6.83 (4.81) | 44.96 | -4.96 (-5.92, -4.00) | 12 | 6 |  |  |
| PHQ8 | Urban | 1465 | 12.74 (4.90) | 7.35 (5.47) | 42.31 | -5.29 (-5.67, -5.10) | 12 | 6 | W = 70020 (df=1) | .065 |
|  | Rural | 107 | 13.47 (4.95) | 7.12 (5.62) | 47.14 | -6.35 (-7.39, -5.30) | 13 | 5 |  |  |

Note: Patients with at least mild baseline symptoms (baseline GAD7/PHQ8 $\geq 5$ ) are included in the analyses.

### Clinically Significant Symptom Improvement (50% or greater reductions in GAD7 and PHQ8)

Of 1074 patients who reported at least moderate anxiety symptoms at baseline (GAD7 ≥ 10), 574 (53.45%) patients showed clinically significant improvement of anxiety symptoms (50% or greater reductions in GAD7). This included 31% of the patients in the moderate category and 21% of patients in the severe category that achieved remission. Among 1097 patients who reported at least moderate depressive symptoms at baseline (PHQ8 ≥ 10), 562 patients (51.23%) were considered to show clinically significant symptoms improvement for depressive symptoms (50% or greater reductions in PHQ8). These values included 37% of moderate, 27% of moderately severe and 13% of severe categories of patients who achieved remission (Figure 1). Of the 1074 patients who reported at least moderate anxiety symptoms at baseline, 724 (67.41%) showed only minimal or mild symptoms at last visit. Of the 1097 patients who reported moderate or greater depressive symptoms at baseline, 685 (62.44%) reported only mild or minimal symptoms at last visit (Table 6).

**Figure 1.**
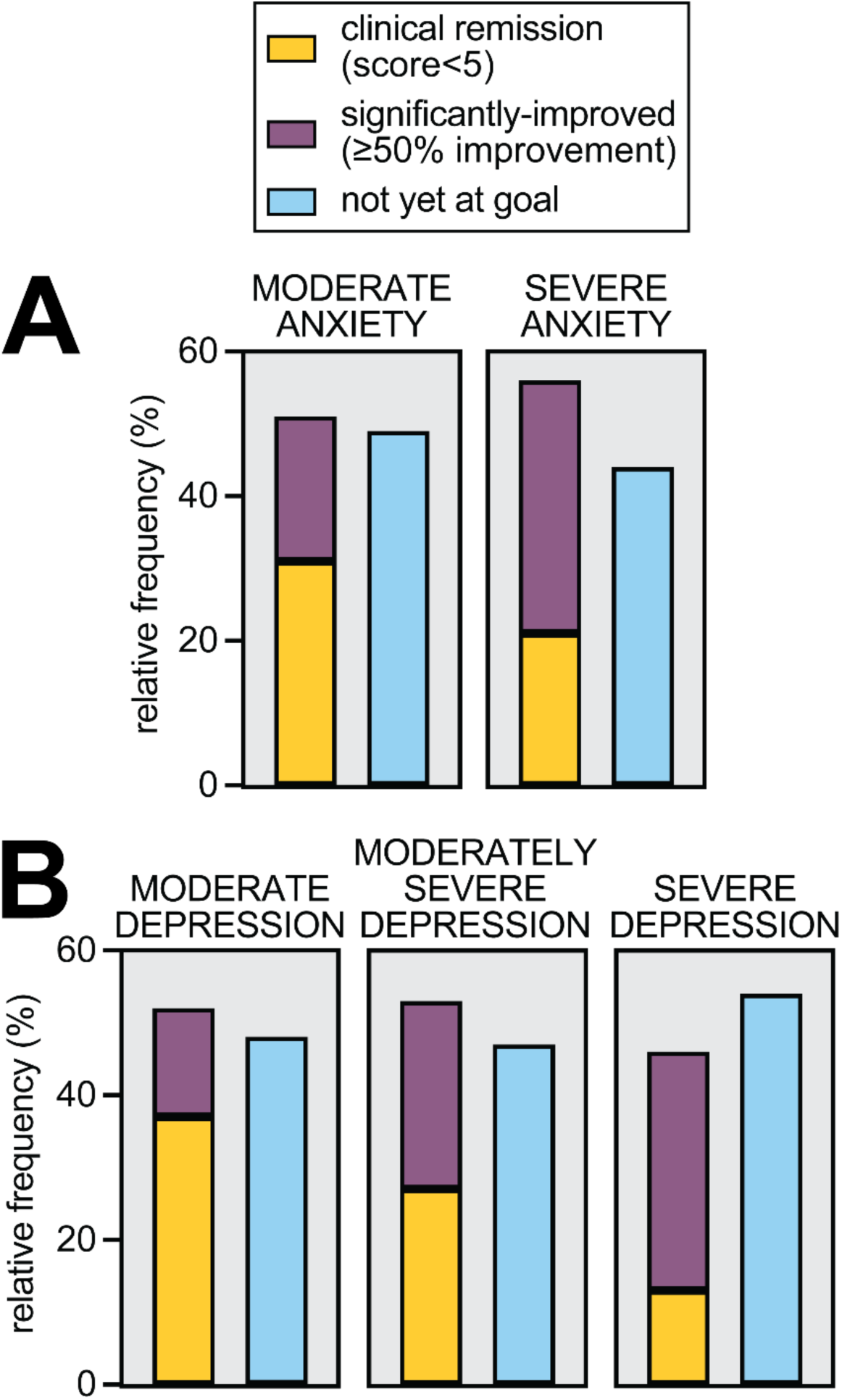
**(A)** Percentage of patients with ≥ 50% improvement in anxiety symptoms, including 31% and 21% respectively reaching clinical remission. **(B)** Percentage of patients with ≥ 50% improvement in depressive symptoms, including 37%, 27%, and 13% respectively reaching clinical remission.

**Table 6.**

| Measure | Responses,<br>n | Patients with 50% or Greater<br>Symptom Improvement,<br>n (%) | Patients with final<br>score of minimal or<br>mild symptoms,<br>n (%) |
| --- | --- | --- | --- |
| GAD7 | 1074 | 574 (53.45%) | 724 (67.41%) |
| PHQ8 | 1097 | 562 (51.23%) | 685 (62.44%) |

Note: Patients with at least moderate baseline symptoms (baseline GAD7/PHQ8 $\geq 10$ ) are included in the analyses. Minimal or mild symptoms = PHQ8/GAD7 $< 10$

#### Baseline Symptom Severity

Among those with at least moderate baseline symptoms, the rates of patients with 50% or greater reductions in either anxiety symptoms (GAD7) or depressive symptoms (PHQ8) did not differ across baseline symptom severity category (*P* = .173, *P*=.341 respectively). This indicates that fractions of patients who showed clinically significant symptom improvement were similar across patients with moderate and severe baseline symptoms (Table 7).

**Table 7.**

| Measure | Baseline Severity | Responses,<br>n (%) | Patients with 50% or Greater<br>Symptom Improvement,<br>n (%) | Chi-squared Test | *P*-value |
| --- | --- | --- | --- | --- | --- |
| GAD7 | Moderate anxiety (10-14) | 541 (50.37%) | 278 (51.38%) | $\chi^2(1, N=1074) = 1.857$ | .173 |
| | Severe anxiety ($\geq 15$) | 533 (49.63%) | 296 (55.53%) | | |
| PHQ8 | Moderate depression (10-14) | 530 (48.31%) | 274 (51.70%) | $\chi^2(2, N=1097) = 2.153$ | .341 |
|  | Moderately severe depression (15-19) | 394 (35.92%) | 208 (52.79%) |  |  |
| | Severe depression ($\geq 20$) | 173 (15.77%) | 80 (46.24%) | | |

Note: Patients with at least moderate baseline symptoms (baseline GAD7/PHQ8 $\geq 10$ ) are included in the analyses.

## Discussion

### Principal results

In this retrospective study, we found that telepsychiatry treatment outcomes for depression and anxiety were statistically and clinically significant. Telepsychiatry treatment led to average reductions of 45.7% for anxiety symptoms and 45.1% for depressive symptoms. Patients with baseline anxiety or depression with severity in the moderate to severe ranges exhibited improved outcomes at the last visit — 67% of patients with anxiety and 62% of patients with depression reported minimal or mild symptoms in response to an average of 103 days of treatment. Further, mean difference between initial and final scores were 42%-45%, which points to robust clinical responses. Interestingly, the extent of improvement was not a function of baseline severity indicating that the efficacy of treatment was comparable among patients that exhibited either moderate or severe symptoms at the start of treatment.

Our findings are consistent with prior research on efficacy of therapeutic alliance in treatment, and provide evidence in support of the notion that therapeutic alliance can be achieved in a telepsychiatry setting. Unique aspects of this psychiatric model include longer visits to allow for the development of effective therapeutic alliance as well as allowing patients to utilize their health insurance which increase access to treatment and reduce the total cost of care. Improving access by reducing cost, reducing delays in seeking treatment, and having confidence in the working relationship with the psychiatrist are important components that improve clinical outcomes in a telepsychiatry platform.

We also found that among individuals utilizing telepsychiatry, rural patients benefitted to the same extent as did urban patients. This finding is promising because it is known that rural patients frequently struggle with access to care due to difficulties with either internet connectivity or geographical constraints [34]. While this study does not address the issue of internet connectivity, our findings argue that in the presence of acceptable internet quality, rural patients were able to access telepsychiatry treatments and showed improvements that were comparable to those seen in their urban counterparts.

Another demographic feature we examined was insurance type. While patients with Medicare insurance in the telepsychiatry practice did exhibit symptom improvements over the course of treatment, the extent of improvement was lower than that observed in patients with commercial insurance or self-pay. This finding is consistent with prior reports that Medicare beneficiaries typically take longer time to respond to treatment, and have more overall disease burden which impacts improvement [35,36].

### Limitations

This study has some limitations. Using data from a private practice utilizing telepsychiatry as the exclusive mode of treatment, we did not have access to a control group that received in-person treatment. This precludes a direct comparison to in-person studies. That being said, we have found that our depression and anxiety outcomes are similar to other such studies [18,37]. A second limitation in this study is that our sample had a comparatively small number of Medicare beneficiaries and a small number of patients older than 65. Due to this limitation, we were unable to determine which of these variables (age, Medicare status, or both) impacted the mean improvement scores. In addition, there were more rural residents who completed both measures, and while they rural residents did not differ in treatment outcomes from urban residents, they may have differed from rural residents who only completed one measure. Another limitation is that other psychiatric conditions which were excluded such as primary substance use disorder, primary psychotic disorder, and primary eating disorder may limit generalizability of the results. As there is considerable comorbidity with other psychiatric illnesses, these treatment outcomes may not be fully representative of all patients who have depression and anxiety. Lastly, although our data are consistent with the establishment of an effective therapeutic alliance, this retrospective analysis lacks quantitative measures of therapeutic alliance. Incorporating measures of therapeutic alliance into routine clinical practice could allow direct evaluation of this relationship in the future.

### Comparison with Prior Work

This study adds to the emerging body of literature that is showing that telepsychiatry can be an effective treatment modality for many mental health conditions [37,38].

## Conclusion

In this large real-world retrospective data analysis of patients utilizing a telepsychiatry platform, we found robust and clinically significant improvement in their depression and anxiety symptoms during the course of treatment. For patients with moderate to severe symptoms of either anxiety or depression, the improvements are similar and confirm that patients with severe illness can be effectively treated in a telepsychiatry practice. This adds to the growing body of medical literature that telepsychiatry can be effective.

## Data Availability

All data produced in the present study are available upon reasonable request to the authors

## Acknowledgements

We would like to thank the psychiatrists at Talkiatry who are transforming psychiatry with accessible, human and responsible care. We also thank Haruka Minami for assistance with statistical analyses and Jared Camins for extensive data support and manuscript review.

## Author Contributions

CP, NK, LK, and KV analyzed the data. CP and GG conceived the study. CP and KV wrote the manuscript.

